# Uncovering dengue serotype-specific transmission, cross-reactivity, and immune profiles from cross-sectional serosurveys

**DOI:** 10.64898/2026.07.23.26358771

**Authors:** Yining Chen, Angkana T. Huang, Hannah Clapham

## Abstract

Dengue remains a growing global health concern, with four co-circulating serotypes (DENV-1 to DENV-4). Cross-sectional serosurveys are essential for inferring past transmission and population immunity, but serotype-specific interpretation is constrained by antibody cross-reactivity across serotypes. Here, we developed a novel modelling framework to jointly infer serotype-specific force of infection (FOI) and cross-reactivity from cross-sectional serosurveys. The model is flexible to data granularity, functioning with binary serostatus alone or combined with quantitative titres. In simulations, the model effectively recovered FOI and cross-reactivity across varying endemicity, sampling designs, and sample age coverage. Applied to annual cross-sectional serosurveys in Vietnam (2013-2017), the model estimated serotype-specific FOI patterns comparable in overall magnitude to analysis requiring supplementary longitudinal post-infection antibody measurements, which are difficult to collect. We further identified higher DENV-3 transmission intensity than previously estimated. Our estimates revealed asymmetric cross-reactivity between infecting and heterologous antibody-response serotypes after primary infections, and high cross-reactivity after post-primary infections. We reconstructed population immune profiles by age, time, serotype, and past infection number, resolving single-serotype and multi-serotype exposure histories not directly distinguishable from observed seroprevalence. Our estimates showed that susceptibility to secondary infection in Vietnam peaked at approximately age 10 for each serotype, and averaged ∼25% for DENV-1 to DENV-3 and ∼30% for DENV-4 among individuals aged 1-30 years. Additionally, incorporating titres enabled individual-level infection-history inference and revealed exposure-history signals in titre profiles despite extensive cross-reactivity. These findings show that improved modelling can substantially expand the information recoverable from cross-sectional serology, strengthening the public-health utility of serosurveys for risk assessment, burden estimation, and vaccination planning.

## INTRODUCTION

Dengue is a major and expanding global public health burden, with reported incidence increasing ten-fold over the past two decades (*1*). Infection can result in outcomes ranging from asymptomatic or mild febrile illness to severe dengue, including dengue haemorrhagic fever and shock syndrome (*1*). Dengue virus (DENV), transmitted primarily by *Aedes* mosquitoes, comprises four antigenically related serotypes (DENV-1, DENV-2, DENV-3, DENV-4) that often co-circulate and induce extensively cross-reactive antibody responses (*2*). Serotype-specific transmission is central to dengue epidemiology and control because immunological interactions among DENV serotypes shape dengue disease risk, epidemic dynamics, and vaccine performance. As short-term cross-protection wanes after primary infection, secondary dengue infections carry increased risk of severe illness through enhancing viral entry into immune cells, which is known as antibody-dependent enhancement (ADE) (*3, 4*). At the population scale, temporal shifts in dominant serotypes are often associated with outbreak peaks, driven by the combined effects of population susceptibility and disease-enhancing immunity (*5*). In addition, dengue vaccines have shown heterogenous efficacy by serotype and pre-vaccination serostatus (*6*), making local serotype circulation and baseline immunity structure critical determinants of setting-specific vaccination effectiveness.

Cross-sectional serosurveys are widely used to monitor population exposure history through presence of anti-DENV antibodies, capturing both reported and unreported infections. However, resolving dengue serotype-specific information is challenging and underexploited. Current neutralizing assays cannot reliably distinguish homotypic antibodies to the infecting serotype from cross-reactive antibodies to other serotypes (*2*). This limits our ability to decipher the complex epidemiological features arising from serotype co-circulation and to distinguish immune profiles with different implications for disease severity. Improved serotype-specific interpretation of serosurveys is therefore critical for translating these data into actionable public health insights for outbreak preparedness, disease burden assessment, and vaccination strategies.

Longitudinal serological cohorts where individuals are tracked after known infection events have provided useful insights into dengue cross-reactivity (*7, 8*), but their applicability to direct serotype-specific interpretation of cross-sectional serosurveys remains limited. These cohorts do not capture virus transmission intensity at the population level, and their short following-up periods, typically less than 3 years, are less informative for the analysis of cross-sectional serosurveys that contain signals of virus transmission dynamics from several years earlier. Moreover, their resource-intensive nature limits spatial and temporal coverage. Recently, a framework was proposed to identify historical infecting pathogens from cross-sectional serology under cross-reactivity (*9*). Nevertheless, it requires titre measurements which are sometimes unavailable. In this method, the transmission dynamics are also estimated separately from between-pathogen cross-reactivity, missing the opportunity to account for infection histories in tandem with cross-reactive antibody responses.

To address this key methodological gap and strengthen the use of cross-sectional serology in dengue control, we developed a cross-reactive catalytic model framework that jointly infers serotype-specific dengue infection risk and cross-reactivity from cross-sectional serosurveys, providing detailed quantification of population immune profiles stratified by serotype and infection number. To resolve the biological ambiguity of cross-sectional antibody profiles, in which different exposure histories can produce similar serological responses, our approach conditions antibody responses on the probabilistic infection risk associated with a participant’s age and sampling date. This framework maintains low data requirements and broad applicability using serostatus data alone. Our model extension, which incorporates antibody titres, further enables individual-level reconstruction of exposure histories and characterization of titre distributions by inferred infection history. Applying our models to cross-sectional serosurveys from Vietnam, we provide context-specific, serotype-resolved evidence on dengue transmission and immunity, enhancing the epidemiological basis for dengue risk assessment and control.

## RESULTS

### Dengue serotype-specific transmission in Vietnam

We analysed the cross-sectional serosurveys from individuals aged 1-30 years, collected annually during 2013-2017 in Ho Chi Minh City (HCMC; 531 samples) and Khanh Hoa (KH; 460 samples) (*10*). Observed antibody titres against dengue serotypes, measured by multiplex protein microarray (PMA), showed extensive cross-reactivity, alongside clear variation across age, time, and location (Figure 1). Titre profiles were often polarized, with most individuals either seronegative to all four serotypes, mainly in younger age groups, or seropositive to all four serotypes. Monotypic and bitypic seropositivity profiles were less frequent and occurred primarily in younger individuals (Supplementary Figure S1). Titres at the assay maximum were concentrated predominantly among individuals older than 15 years in HCMC, with no apparent clustering by sampling year. In KH, maximum titres were observed across age groups older than 5 years. Notably, among children aged 5–10 years in KH, these maximum titres were mostly observed in samples collected during 2016– 2017 across all four serotypes.

**Figure 1.**
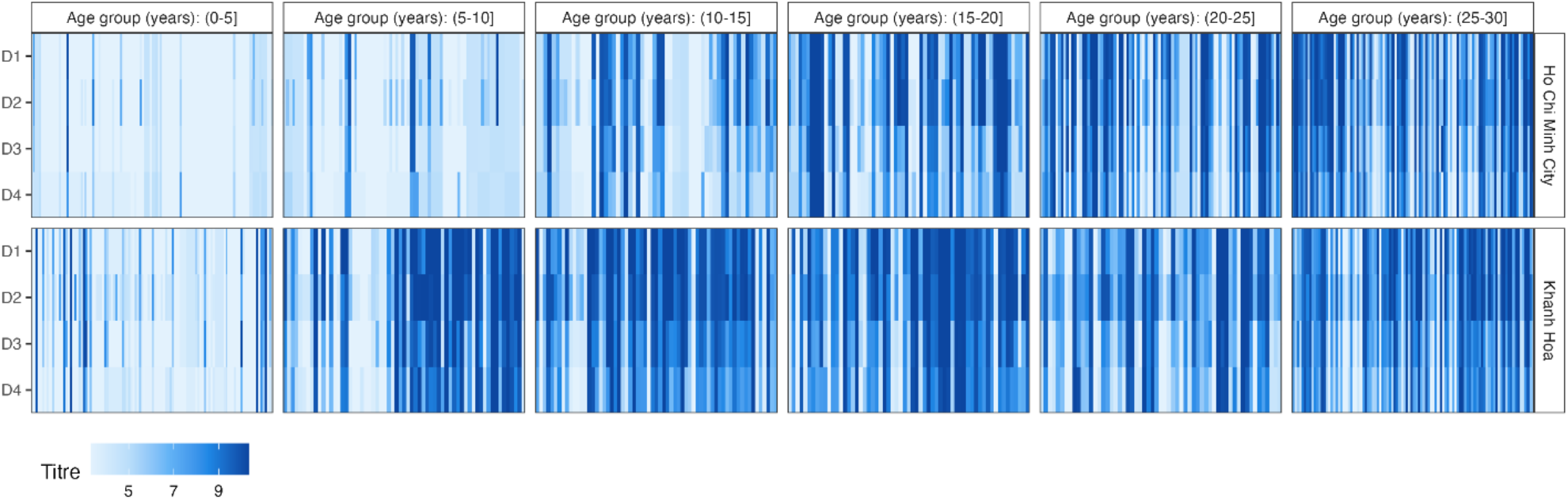
Individual-level titre profiles. Each column represents one individual. Within each panel, individuals are ordered along the x-axis according to sampling time, from 2013 to 2017. A titre value of 5 was used as the seropositivity threshold, based on calibration against negative-control samples (*10*).

To disentangle the serotype-specific epidemiological signals from these highly cross-reactive antibody response profiles, we developed two cross-reactive catalytic models: a serostatus-based model using binary serostatus data, and a titre-incorporated model additionally using quantitative antibody titres. We assumed up to 4 times of infection (once with each of the four serotypes), life-long immune protection against homotypic infection, and short-term cross protection against heterotypic infection. Based on the estimates of force of infection (FOI), defined as the risk of infection per unit time among susceptible individuals, our two models estimated that the four dengue serotypes co-circulated at generally comparable magnitude in HCMC and KH from the 1980s to 2017, with moderate temporal fluctuation and slightly lower circulation of DENV-4 (Figure 2A, Supplementary Figure S2). Using only cross-sectional serosurveys, our models inferred an overall FOI of generally similar magnitude to previous estimates that were supplemented with additional longitudinal measurements of antibody responses following infection (Supplementary Figure S2) (*10*). Both our estimates and previous estimates showed a pronounced FOI peak in KH in 2016, consistent with the sharp increase in titre values among children aged 5–10 years. Despite this overall similarity, our serotype-specific estimates further identified DENV-3 as a predominant serotype with higher transmission intensity than previously reported.

**Figure 2.**
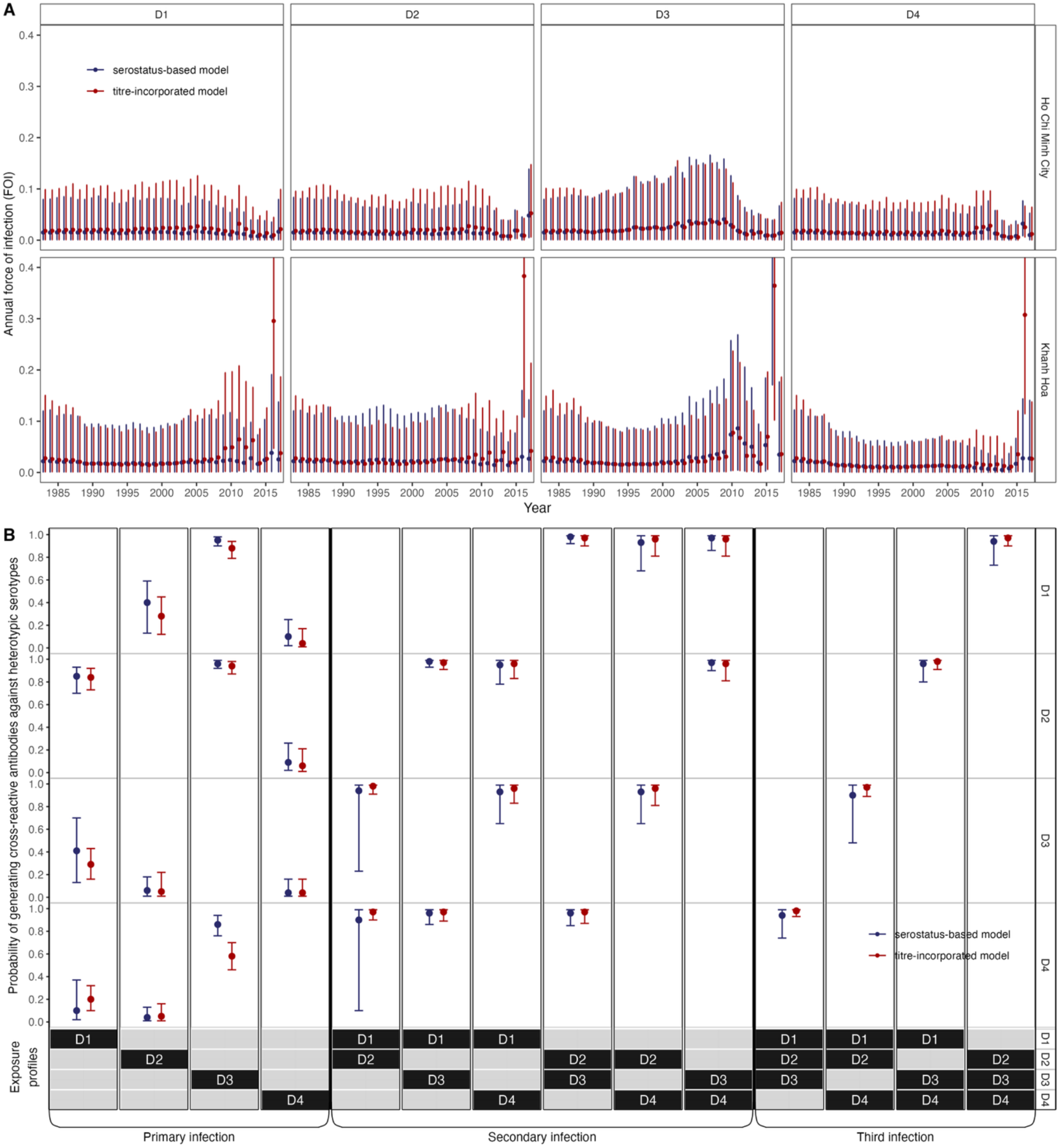
The estimated annual force of infection for 4 serotypes in Ho Chi Minh City and Khanh Hoa (A), and the estimates of cross-reactive probabilities (B). The serostatus-based model was fitted to binary serostatus data, whereas the titre-incorporated model additionally incorporated quantitative antibody titres. The points refer to the estimated median. The error bars refer to the 95% credible intervals. D1, D2, D3, and D4 denote DENV-1, DENV-2, DENV-3, and DENV-4, respectively. A. See Supplementary Figure. S2 for a side-by-side comparison of FOI estimates across four serotypes. B. Each column represents a distinct exposure profile, with the combination of infecting serotypes indicated by black cells at the bottom annotation.

### Dengue cross-reactivity pattern

Our models used probabilities of cross-reactive responses to non-infecting serotypes to quantify the proportion of seropositivity attributable to cross-reactivity. After primary infections, we estimated an overall asymmetric cross-reactivity pattern (Figure 2B). DENV-3 infections have a high estimated probability of generating antibodies cross-reactive to other serotypes. In contrast, primary infections of other serotypes had a relatively low probability of generating antibodies cross-reactive to DENV-3. Similarly, DENV-1 infections were estimated to frequently generate cross-reactive antibodies against DENV-2, whereas DENV-2 infections were less likely to generate cross-reactive antibodies against DENV-1. After post-primary infections, the estimated probability of generating cross-reactive antibodies against all heterotypic serotypes was high, at approximately 0.96-0.98 (Figure 2B).

### Reconstructed immune profiles and their divergence from seroprevalence profiles

From our estimated serotype-specific exposure histories, we reconstructed population immune profiles assuming lifelong homotypic immunity after infection. Consistent with the seroprevalence trends, the proportion of fully naïve individuals, who are susceptible to primary dengue infection, declined with age, and reached ∼5% at age of 30 years old in both locations (Figure 3A). For temporal comparisons, estimates were averaged over individuals aged 1–30 years. This age-averaged proportion fluctuated around 26% in both locations during 2000-2017, except for a decline in KH after 2016, which was driven by the high estimated FOI in 2016 (Supplementary Figure. S3).

**Figure 3.**
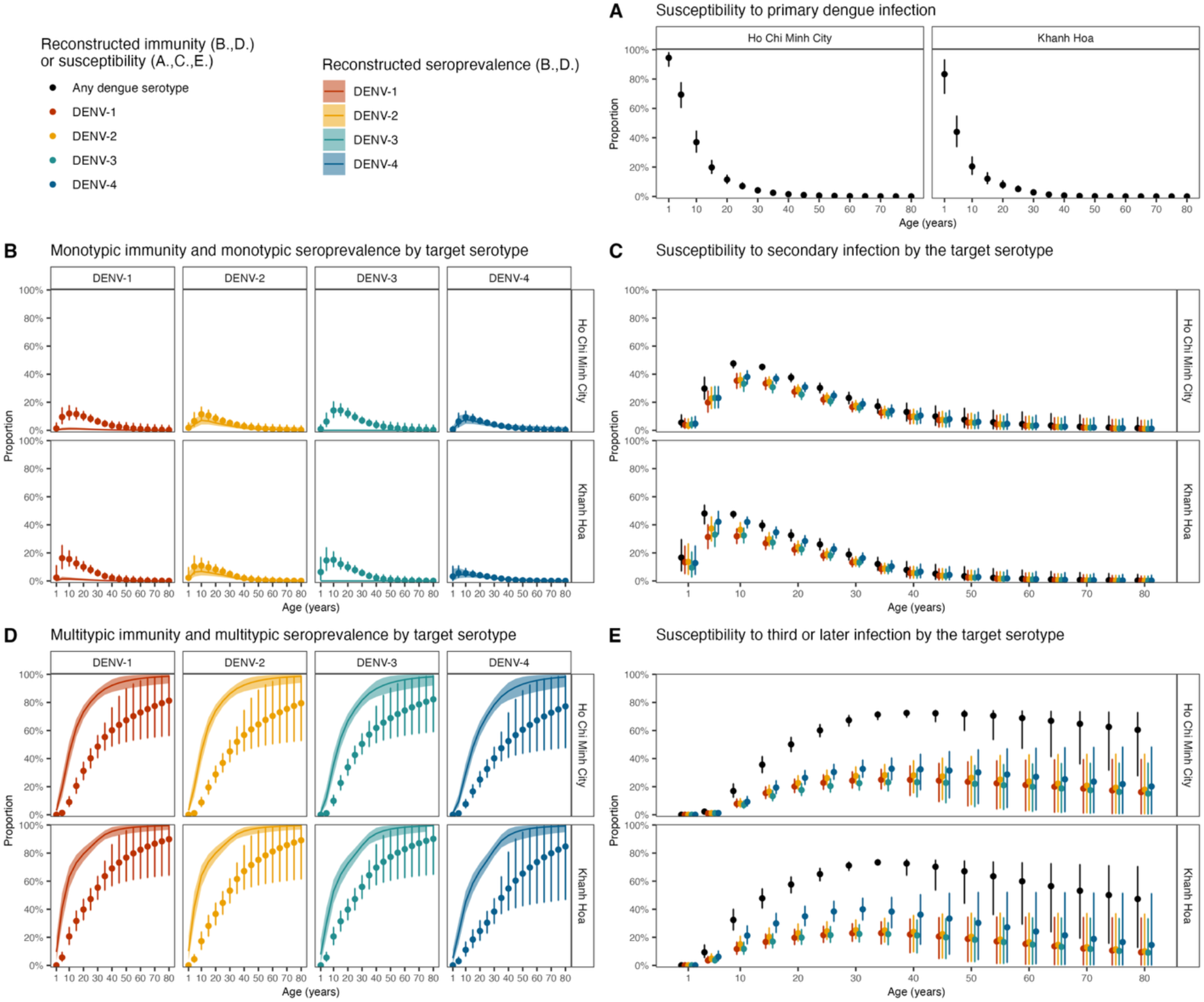
Neglecting cross-reactivity obscures the distinction between single- and multi-serotype exposure histories reflected in seroprevalence. The panels show reconstructed population immunity and susceptibility from the titre-incorporated cross-reactive model by age for the 2015 population, selected as a representative recent year. The time-varying estimates averaged over individuals aged 1-30 years (*11*) are shown in Supplementary Figure. S3, and estimates from the serostatus-based model are shown in Supplementary Figures. S4-S5. Points indicate posterior medians of reconstructed immunity or susceptibility, and error bars indicate 95% credible intervals. In B and D, lines indicate reconstructed seroprevalence medians, and shaded areas indicate 95% credible intervals.

Serotype-specific seroprevalence, which reflects both historical infection and cross-reactive seropositivity, captured broad exposure patterns, but model-based reconstruction was needed to distinguish monotypic from multitypic immunity to each target serotype, defined respectively as prior exposure only to the target serotype or to the target serotype plus at least one other serotype. Monotypic immunity was markedly higher than the corresponding serotype-specific monotypic seroprevalence for DENV-1 and DENV-3, slightly higher for DENV-2, and similar for DENV-4 (Figure 3B, Supplementary Figure. S3). Susceptibility to secondary infection by a given serotype, defined as prior exposure to one heterologous serotype only, concentrated in younger age groups, peaking at 5-10 years in HCMC and 10-15 years in KH (Figure 3C). Across the four serotypes, this susceptibility was slightly higher for DENV-4 due to its lower estimated circulation intensity as shown in FOI. The age-averaged susceptibility remained broadly stable over time in both locations, at approximately 25% for DENV-1, DENV-2, DENV-3, and approximately 30% for DENV-4 (Supplementary Figure. S3).

Serotype-specific multitypic immunity was considerably lower than the corresponding multitypic seroprevalence for all four serotypes (Figure 3D, Supplementary Figure. S3). Consistent with the age-related accumulation of exposure apparent in the seroprevalence data, this multitypic immunity, as well as the proportion remaining susceptible to third or later infection, increased with age (Figure 3D–E). During 2000–2017, the age-averaged proportion remaining susceptible to third or later infection by each of the four serotypes fluctuated around 15% in both locations (Supplementary Figure. S3).

### Exposure-history signals in dengue antibody titre profiles despite broad cross-reactivity

Further incorporation of titres (titre-incorporated cross-reactive model) enabled us to decompose antibody titres into additive infection-number effects and response-specific boosts. In this model, titres increase with successive infections (infection-number effects) through non-negative increments for previously exposed, and separately, non-exposed serotypes. Titres to exposed serotypes were assumed to reach the assay maximum after the third infection. Response-specific titre boosts captured elevation attributable to direct responses to infecting serotypes or cross-reactive responses to heterologous serotypes, and were estimated separately for each serotype. The model also allowed for additional titre boosts among dengue-naïve and monotypically-infected individuals to capture potential non-dengue antigen effects on titre distributions. The titre-incorporated model inferred individual-level infection histories, with estimated titre distributions closely reproducing the observed titre patterns across exposure profiles (Figure 4).

**Figure 4.**
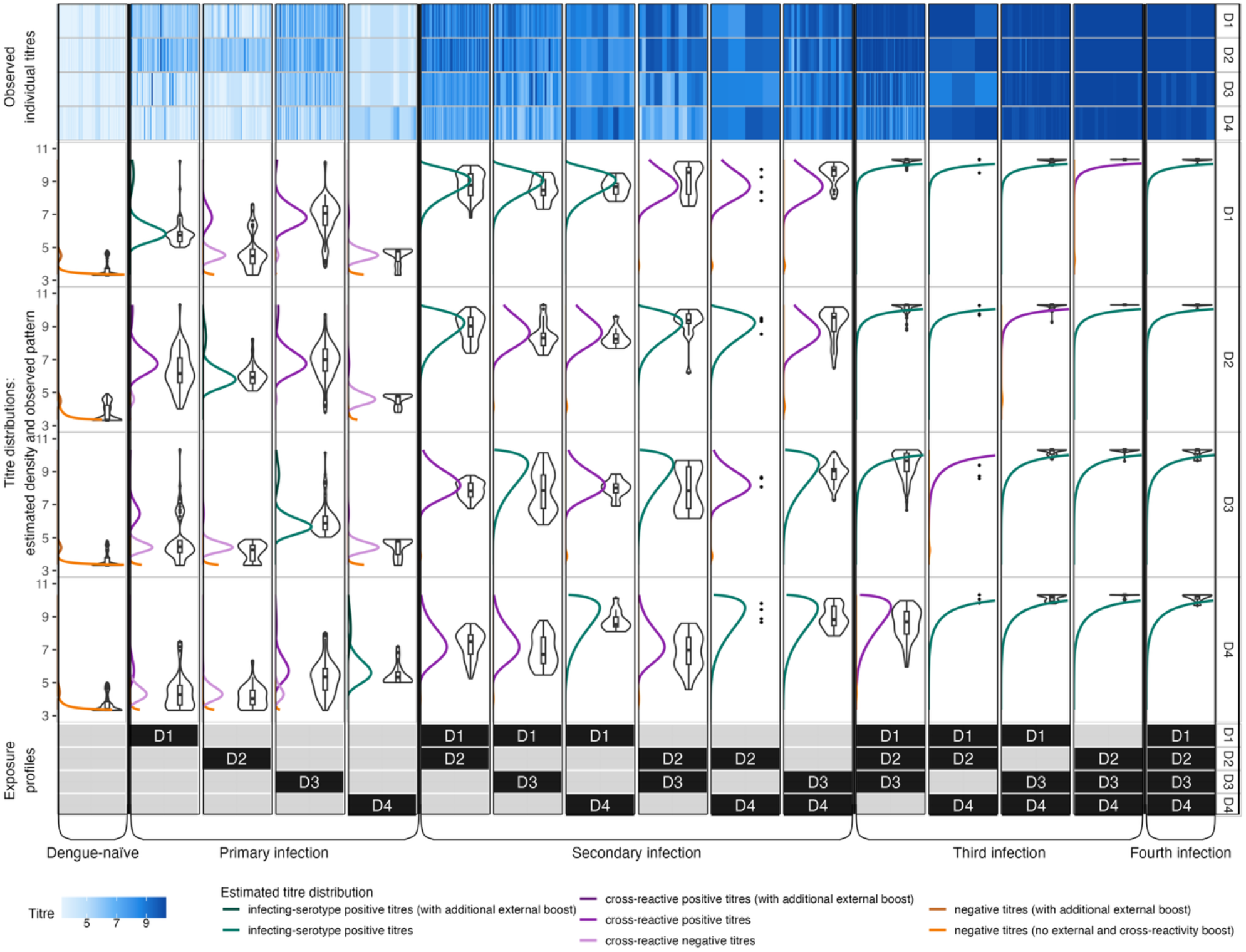
Titre distributions by estimated infection history. Each column represents a distinct exposure profile, as defined in the bottom annotation, with black cells indicating the infecting serotypes included in that profile. The upper panels show observed individual-level titre values, and the middle panels compare model-estimated and observed titre distributions across estimated infection histories. In the middle panels, coloured lines indicate model-estimated titre distributions, while black violin plots with embedded box plots summarize observed titre values from the individuals shown in the corresponding upper panels. Points above the box plots indicate outliers. For exposure profiles represented by fewer than five individuals, observed titre values are shown as points. Posterior estimates of all titre parameters are provided in Supplementary Table S1.

The immune response after primary infections with different serotypes revealed distinct titre combinations across the four serotypes (Figure 4). After primary DENV-1 infections, mean titres were higher for DENV-1 and DENV-2 and lower for DENV-3 and DENV-4. After primary DENV-2 infections, DENV-2 titres predominated, followed by a moderate rise in DENV-1 and negligible responses to DENV-3 and DENV-4. Primary DENV-3 infection showed wider elevation across DENV-1, DENV-2, and DENV-3, with a modest increase for DENV-4 that remained comparatively low. Primary DENV-4 infection was characterized by elevated DENV-4 titres, while titres for the other three serotypes remained low. Post-primary titre profiles showed infection-number gradients despite extensive cross-reactivity (Figure 4). This pattern was evident in both the estimated and observed titre distributions, with titres for both infecting and non-infecting serotypes increasing with reconstructed infection number.

The titre-boost component in dengue-naïve and primary-infected individuals was consistent with exposure to non-dengue antigens. Individuals assigned to this sub-population had significantly higher titres (*p* < 0.001) against other flavivirus antigens (St. Louis encephalitis Virus, Zika Virus, and West Nile Virus) measured on the same platform (information not used in the modelling process, Supplementary Figure. S6). The estimated proportion with this additional boost was highest in HCMC during 2017 (50%), and was also relatively high in KH during 2015 (17%) and 2016 (12%), coinciding with the documented 2015-2016 Zika virus outbreak in Vietnam (*12*).

### Comparable inference from serostatus-based and titre-incorporated cross-reactive models

The serostatus-based model, despite using binary serostatus data alone, recovered serotype-specific FOI and cross-reactivity estimates comparable to those from the titre-incorporated model (Figure 2). Compared with the serostatus-based model, the titre-incorporated model estimated slightly higher DENV-1 and DENV-2 transmission in both locations. The main difference occurred in KH in 2016, where both models captured a marked transmission peak but interpreted the underlying serological pattern differently. The serostatus-based model interpreted the 2016 shift toward seropositivity against three or four serotypes among children aged 5–10 years in KH primarily as recent DENV-3 infection. This was consistent with the high estimated probability of cross-reactive responses following DENV-3 infection. By incorporating titre magnitudes, the titre-incorporated model attributed more of the multitypic high-titre profiles near the assay maximum to multiple infections, estimating increased FOI across the four serotypes. FOI uncertainty was comparable between models, whereas incorporating titre values substantially reduced uncertainty in cross-reactivity estimates, especially for post-primary infections, and enabled individual-level reconstruction of infection histories. Together, these comparisons showed that the serostatus-based model maintained comparable serotype-specific inference when quantitative titre values were reduced to binary serostatus (the form of data more commonly available).

### Model applicability across varying endemicity, sampling strategies, and age coverage

We next used simulation studies to evaluate the serostatus-based model under increasing constraints in data availability and resolution. Model performance was assessed under endemic settings with low- and high FOI across scenarios varying in data structure, sampling frequency, and age coverage. Data structures included complete data, in which serotype-specific serostatus was measured for all samples as in Vietnam serosurveys (*10*), and subset-tested data, in which serotype-specific serostatus was measured only in a subset of dengue immunoglobulin G (IgG)-positive samples, as commonly used in Singapore serosurveys (*13, 14*). This subset-tested design is lower-cost and widely feasible in practice. We also varied sampling frequency (annual consecutive sampling versus 4-year periodic sampling) and age coverage (ages 1–75 years versus exclusion of younger or older ages). Across these scenarios, the model successfully recovered FOI dynamics and cross-reactivity patterns, with performance for these two inference targets depending on different factors (Figure 5, Supplementary Figures. S7-S11).

**Figure 5.**
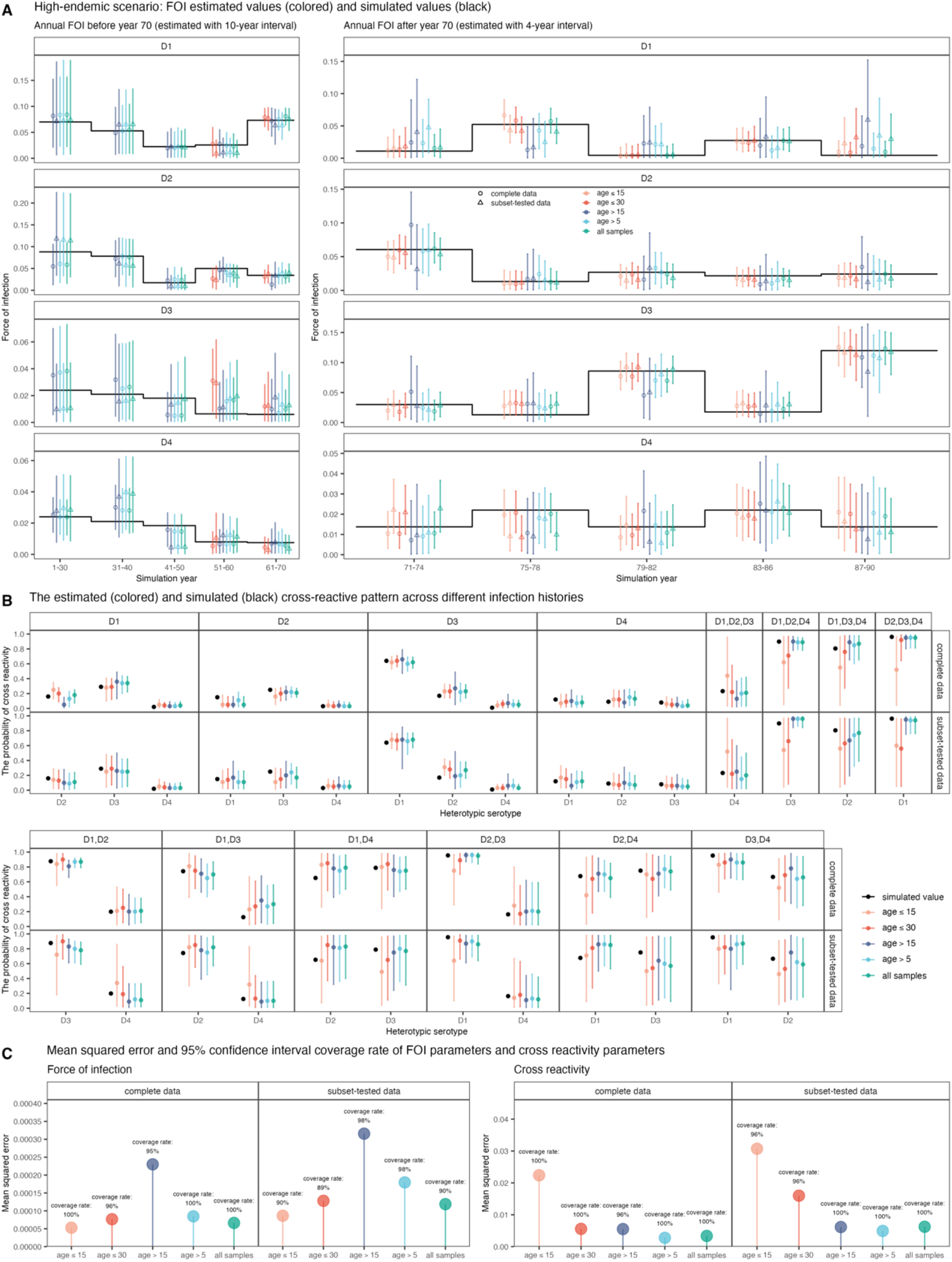
The model performance in simulation study under periodic sampling frequency in high-endemic setting. See Supplementary Figures. S7-S11 for model performance under other sampling frequency and in low-endemic setting. A., B. The simulated values and estimated values of FOI during simulation years 1-90 and cross-reactivity probabilities. The coloured points refer to the posterior median estimation. The coloured error bars refer to the estimated 95% credible intervals (CrI). The black lines and points are the simulated values. C. The mean squared error between simulated values and estimated values and the 95% CrI coverage rate. The 95% CrI coverage rate is calculated as the proportion of parameters with simulated values covered by the estimated 95% CrI.

The model ability to infer FOI depended mainly on the complexity of mixed exposure histories. In low-endemic setting, where all four serotypes were present but circulated at low intensity, subset-tested data sometimes performed slightly better than complete data in FOI inference, likely because non-serotype-specific IgG serostatus provided stable information on overall dengue exposure, which the model could infer more easily than serotype-specific exposure allocation, helping to anchor serotype-specific FOI estimates. In high-endemic settings, the two data structures performed similarly. Restricting samples to younger ages (age ≤ 15, age ≤ 30) improved FOI estimation in several settings, in line with the simpler exposure histories and closer link to recent transmission in younger individuals, although this came at the cost of reduced information on earlier transmission. In contrast, using samples excluding younger ages (age > 5, age > 15) substantially reduced accuracy across all scenarios, with larger losses as more young age groups were omitted.

The model ability to infer cross-reactivity depended mainly on the amount and diversity of serotype-specific information in the sample. Complete data generally performed better than subset-tested data in cross-reactivity inference, as measuring serotype-specific serostatus in all samples may provide richer information on cross-reactive relationships between serotypes. Restricting samples to younger ages (age ≤ 15, age ≤ 30) reduced accuracy and precision, likely attributable to fewer represented exposure histories. In addition, the estimate uncertainty in post-primary cross-reactivity pattern was high in both endemic settings, consistent with patterns observed in the Vietnam data analysis. This effect was more pronounced under low endemicity, where fewer post-primary infections may reduce the statistical power for precise estimation.

In additional simulated scenarios, the model consistently recovered FOI and cross-reactivity patterns when simulated cross-reactivity values were randomly selected or drawn from posterior estimates from the Vietnam data analysis, indicating robustness of model performance to different assumed cross-reactivity structures (Supplementary Figure. S12). Using misspecified priors on cross-protection duration (Supplementary Equations (Eqs.) S1-S2) caused a slight decline in FOI performance, with larger variability in posterior median FOI estimates under high-endemic setting (Supplementary Figures. S13-S16).

### Sensitivity of model inference

In the Vietnam data analysis, posterior estimates were largely robust to the informative prior of cross-protection duration. Compared with the main analysis, which used a prior centred at 1.8 years, alternative priors (no cross-protection; 1-year; 3-year) produced similar FOI and cross-reactivity estimates. Notable differences were limited to the reduced FOI estimates in KH during 2016 under shorter cross-protection duration in both models, and minor fluctuations in the estimated probability of generating antibodies cross-reactive to DENV-4 following DENV-3 infection in the serostatus-based model (Supplementary Figures. S17–S20).

## DISCUSSION

We developed a cross-reactive catalytic modelling framework that enables joint inference of serotype-specific dengue transmission and cross-reactivity from cross-sectional serosurveys. Our findings demonstrated that dengue antibody response profiles contained exposure-history signals despite extensive cross-reactivity. The results confirm previous virological surveillance evidence of serotype co-circulation in Vietnam (*15*), showing that all four dengue serotypes sustained broadly comparable transmission in southern and central Vietnam over recent decades, and extend this evidence by quantifying the underlying transmission intensity of each serotype, beyond their observed distribution among reported cases. We estimated an asymmetric cross-reactivity pattern after primary dengue infection, and a high probability of cross-reactivity to heterotypic serotypes (> 95%) after post-primary infection. By reconstructing serotype- and infection-number–specific population immune profiles, we showed that the population at risk of secondary infection in Vietnam peaked around 10 years of age for each of the four serotypes and was highest for DENV-4. This profile is not directly captured by seroprevalence, which cannot explicitly distinguish single from multiple past exposures, and goes beyond non-serotype-specific FOI analyses by resolving secondary-infection susceptibility separately for each target serotype.

Our framework substantially expands the epidemiological information recoverable from cross-sectional serology and remained applicable across common serosurvey designs with varying sampling frequency, age coverage, and serotype testing schemes, as evaluated in the simulation study. These gains in inferential scope do not require increased data collection in serosurveys that use serotype-specific assays, as the framework can incorporate titre values when available but also supports reliable inference from binary serostatus data alone. Comparison with previous analyses illustrates this data-efficient inference. Using only cross-sectional serological data, our overall FOI estimates were broadly consistent with previous reports, which relied on additional longitudinal data following up individuals after infections and titres against other measured flaviviruses to make pre-modelling classification of primary infections and identify the corresponding infecting serotype (*10*). Our framework avoids this pre-classification step, jointly estimates past infection status and FOI, allowing uncertainty in both components to be captured within the model. At the serotype level, we estimated a higher transmission intensity for DENV-3. Together with our estimated high probability of cross-reactive responses following DENV-3 infection, this explains the high levels of multitypic seropositivity observed in young children. This difference from the earlier reported estimates may arise from their reliance on the longitudinal cohort with a small proportion of DENV-3 infection (13%, *N* = 19) (*8*), which may have identified more DENV-1 and DENV-2 than DENV-3 infections in their pre-modelling classification.

Our reconstructed serotype- and infection-number-specific immune profiles provide a missing link between population-level historical exposure and observed patterns of dengue disease severity. Dengue clinical outcomes are closely related to exposure history: secondary dengue infections are associated with higher illness severity through ADE, whereas third and later infections are generally less symptomatic. Our reconstructed age pattern of secondary-infection susceptibility was consistent with the surveillance data from southern Vietnam, where most severe cases occurred among children aged 5-9 and 10-14 years (*15*). The observed age distribution of severe dengue likely reflects the joint effects of infection age patterns, age-related clinical risk, serotype-specific clinical presentation, and the population immune landscape. This immune landscape is rarely observed directly, making it difficult to interpret age- and serotype-specific disease risk without quantifying the corresponding numbers of primary and secondary infections. Our reconstructed immune profiles therefore provide a useful foundation for interpreting shifts in disease severity and for enhancing context-specific outbreak preparedness and disease burden assessment.

The serotype-specific FOI estimates also complemented case-based surveillance by revealing temporal transmission dynamics that may be masked by reporting variability and pre-existing immunity. In HCMC, a general decline in transmission was estimated in this study across all four serotypes after 2009, followed by a modest rebound during 2016-2017, consistent with the notified case trend. In Khanh Hoa, our estimated FOI revealed larger temporal fluctuations with frequent shifts in predominant serotypes, whereas notified dengue cases from the surveillance system also showed fluctuating patterns. The cases revealed major peaks in 2005, 2010, 2012-2013, and 2015 (*10*), some of which aligned with our FOI estimates (e.g., 2012-2013 and 2015). Correlation between case peaks and serotype replacement events was observed, such as the rise of DENV-3 in 2010 and 2015, and the resurgence of DENV-1 in 2012-2013. Meanwhile, differences were observed in the timing of peaks between case dynamics and FOI estimates, due to the complex interplay between underlying transmission dynamics, changes in population susceptibility, and year-to-year variation in case reporting rates.

This study recovers antibody-response patterns and provides novel estimates of dengue cross-reactivity from cross-sectional serology. In some primary-infection profiles, mean titres to heterotypic serotypes among individuals with positive cross-reactive responses exceeded mean titres to the inferred infecting serotype; for example, DENV-1 and DENV-2 titres were higher than DENV-3 titres after inferred primary DENV-3 infection. This is congruent with patterns observed in longitudinal dengue studies showing higher DENV-1 and DENV-2 than DENV-3 titres two years after primary DENV-3 infection (*7*). These patterns are serological plausible, because measured antibody reactivity reflects binding to assay antigens and may be shaped not only by infecting serotype but also by shared epitopes, antigen presentation, and antigen-specific assay sensitivity. Consistent with immune-response accumulation and broad flavivirus cross-reactivity, titres against St. Louis encephalitis Virus, Zika Virus, and West Nile Virus also increased with inferred dengue infection number (*p* for trend < 0.0001 ; Supplementary Figure. S21). Our estimated asymmetric cross-reactivity following dengue primary infections has also been reported in another antigenically related virus system, between seasonal influenza A/H1N1 and pandemic influenza A/H1N1 viruses (*16*). Because serological cross-reactivity may vary with antigenic distance between circulating strains and evolve with dengue virus phylogeny (*17, 18*), our estimates of time-constant cross-reactivity matrix, informed by the overall cross-reactive proportion within our sample, reveal a time-averaged summary of the dominant cross-reactivity pattern during the study period. The consistency of FOI and cross-reactivity estimates between the titre-incorporated and serostatus-based models supports the robustness of these epidemiological inferences, despite the reliance of the titre-incorporated model on titre-distribution assumptions. Future work will test the generalizability of inferred titre profiles and cross-reactivity patterns across serosurveys using other assays. More broadly, our modelling framework provides a feasible basis for further extensions that incorporate the complexities of temporally dynamic serological cross-reactivity.

One limitation of this study is that we did not explicitly model the cross-interactions between dengue viruses and other flaviviruses. However, we did consider this potential confounding on titre values by estimating a sub-population with elevated antibody responses. Furthermore, the computational complexity increases exponentially with the number of antigens considered. For a given serostatus combination, the number of possible infection history scenarios to evaluate is 2^|*T*|^ − 1, where |*T*| is the number of seropositive antigens. For the four dengue serotypes studied here, this computational burden remains manageable. But applications to larger antigen panels may require computational approximations or constraints on plausible infection histories. In the titre-incorporated model, we did not use measurements of responses to non-dengue flaviviruses or infer their transmission, but instead represented their potential impact as an overall effect on dengue titre distributions, providing a practical way to reduce computational complexity.

In conclusion, we developed a catalytic modelling framework that incorporates antibody cross-reactivity, extending conventional inference from cross-sectional serology to jointly estimate serotype-specific dengue transmission and cross-reactivity patterns, thereby to reconstruct population immune profiles. By extracting richer epidemiological information from standard cross-sectional serosurveys, the framework greatly enhances the utility and cost-efficiency of both existing datasets for retrospective analysis and future surveillance efforts. This study provides a novel quantification of dengue cross-reactivity and exposure-history-specific titre distributions, offering new insights into long-term serological interactions. The high-resolution estimates of population immunity and susceptibility in Vietnam help transform routine surveillance into a quantitative evidence base for tailoring public health interventions to local epidemic risk.

## MATERIALS AND METHODS

### Study design

We developed a cross-reactive catalytic model framework to jointly estimate serotype-specific FOI and cross-reactivity from cross-sectional serosurveys. The model is applicable for using binary serostatus alone (serostatus-based model), or additionally incorporating quantitative antibody titre values (titre-incorporated model), which enables individual-level inference of infection history. We evaluated the performance of serostatus-based model in simulation study under low-endemic and high-endemic settings, and across serosurvey scenarios varying in data structure, sampling frequency, and age coverage (Table 1). These scenarios represent common real-world designs, such as the Singapore serosurveys collected every four years among individuals older than 15 years with serotype-specific serostatus measured in a subset of IgG-positive samples (*13, 14*), and the Vietnam serosurveys collected annually among individuals younger than 30 years with serotype-specific serostatus available for all samples (*10*).

**Table 1.**
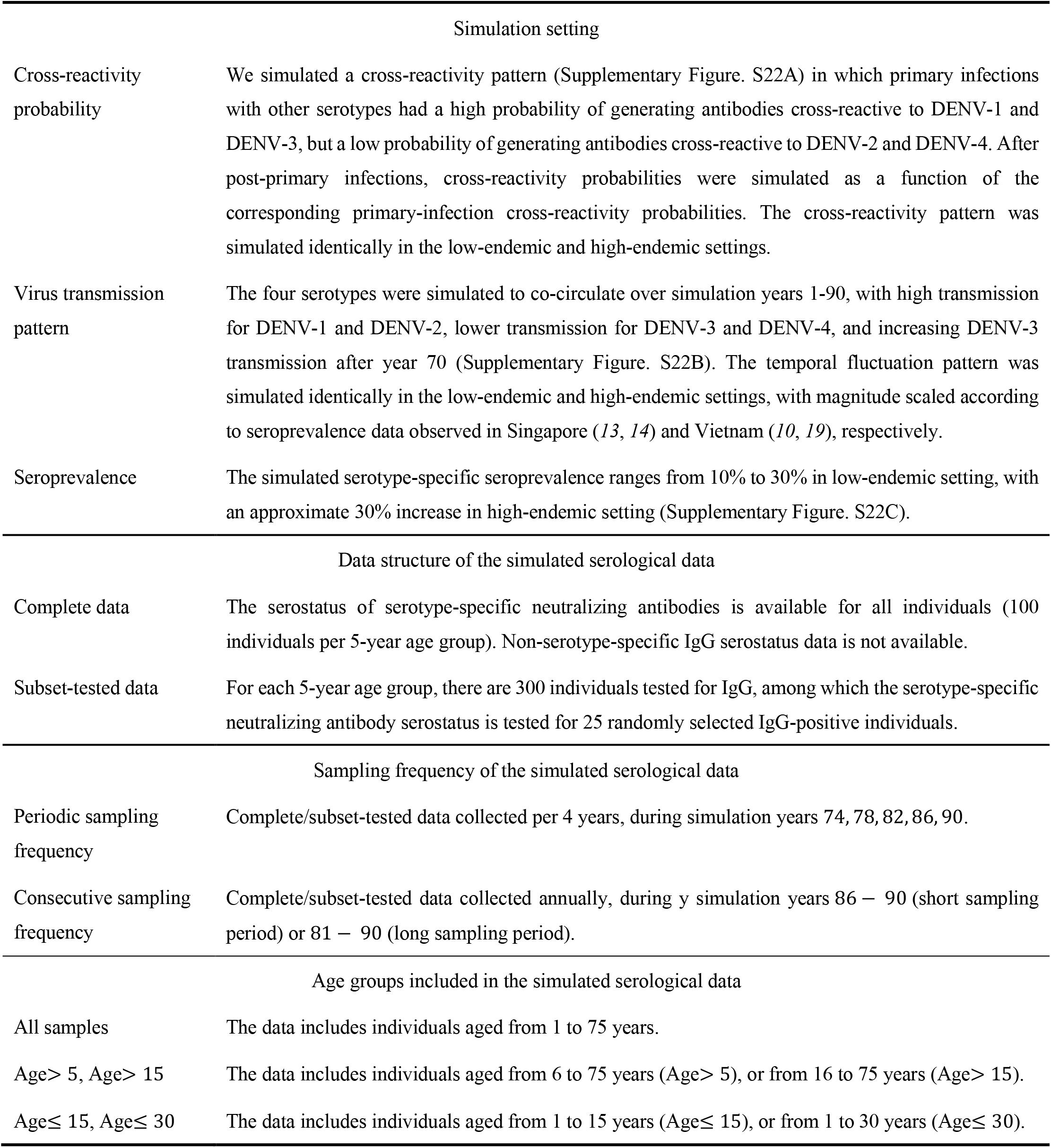
Simulation setting and simulated serological data.

We used the cross-sectional serological data obtained from a serum bank in Vietnam as described previously (*10*). In brief, the analysed residual serum samples were collected annually during 2013-2017 from HCMC (531 samples) and KH (460 samples) respectively. The sample age was from 1 to 30 years old. PMA was used to measure antibody titres against multiple flavivirus antigens, including DENV-1 to DENV-4, St. Louis encephalitis Virus, Zika Virus, and West Nile Virus. Seropositivity was defined as a PMA titre value higher than 5 based on a negative-control calibration (*10*).

### General framework of serostatus-based cross-reactive model

The data observation for the serotype-specific antibody serostatus is defined as *T* ⊆ {D1,D2,D3,D4}, referring to the set of serotypes for which an individual is classified as seropositive. Let ℋ ⊆ {D1,D2,D3,D4} denote a latent variable infection history, referring to the set of infecting serotypes. Let *ϕ*_ℋ,*n*_ = Pr(*n* ∈ *T*|ℋ) refer to the probability of generating positive cross-reactive antibodies against serotype *n* given infection history ℋ (*n* ∉ ℋ).

Under the basic scenario that ignores antibody decay, the model assumes that at least one serotype with positive antibodies is the infecting serotype. The likelihood of serostatus observation *T* = *τ* is calculated as 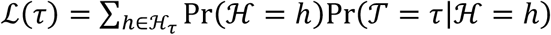 where ℋ *τ* = {ℋ ⊆ *τ*|ℋ ≠ ∅ if *τ* ≠ ∅}. Given the presence of cross-reactivity, ℒ(*τ*) can be expanded as below.

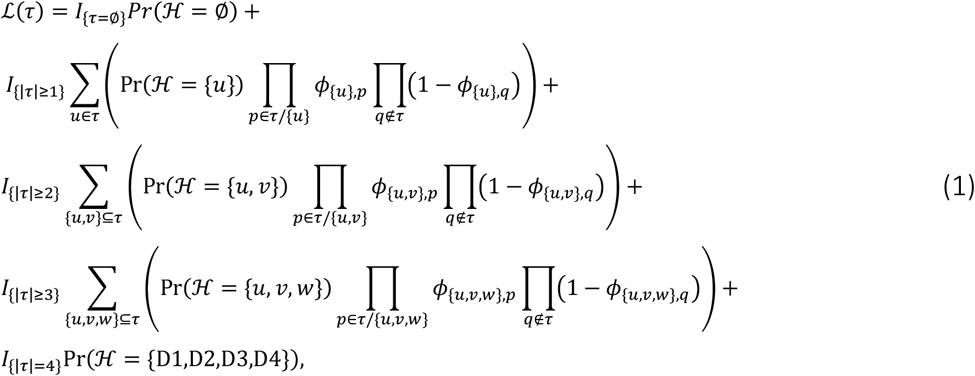

where |*τ*| is the the number of elements in set *τ*, i.e., the number of serotypes to which an individual has a response, *I* is the indicator function which equals to 1 when the condition in the subscript is satisfied and equals to 0 otherwise. Examples of Model (1) are available in Supplementary Eqs. S4-S6.

Model (1) incorporates cross reactivity between dengue serotypes through a flexible structure, where the mathematical form of Pr(ℋ) and *ϕ* can be specified based on different modelling assumptions. The basic catalytic model with no cross-reactivity emerges as a special model of Model (1) with ℋ_*τ*_ = {*τ*} and *ϕ*_ℋ,*n*_ = 0 (details in Supplementary Eq. S7).

In this study, we derived Pr(ℋ) under the assumptions of a maximum of four infections (once with each of the four serotypes), no loss of seropositivity following seroconversion, life-long immune homotypic protection, and short-term heterotypic cross protection. During the cross-protection period, individuals are assumed to be protected from reinfection, and no boosting of antibody titres occurs. We conduct sensitivity analysis to evaluate the impact of cross-protection assumptions on model estimates and model performance.

The probability of no prior infection (ℋ = ∅) for individuals aged *A* during year *T* is calculated as

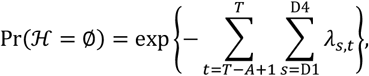

where λ_*S*,*T*_ refers to the annual FOI of serotype *S* during year *T*.

For primary infections (ℋ = {*m*}), Pr(ℋ = {*m*}) is calculated by partitioning on whether infection occurs before or after *T* − *l*, where *l* refers to the short-term cross-protection duration.

With 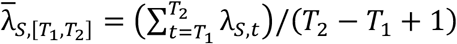 referring to the average annual FOI of serotype *S* from year *T*_1_ to *T*_2_, we denote the average annual FOI during lifetime up to year *T* as 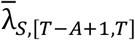, before *T* − *l* as 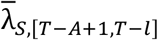, and from *T* − *l* to *T* as 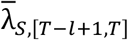. Pr(ℋ = {*m*}) is derived as follows, which nests the general catalytic model when *l* = 0, and satisfies Pr(ℋ = {*m*},infection before *T* − *l*) = 0 for *A* ≤ *l* (details in Supplementary Eqs. S8-S12).

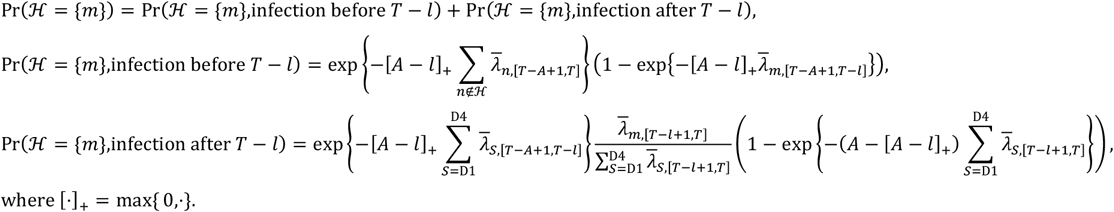

For post-primary infections (|ℋ| > 1), Pr(ℋ) is derived under a simplified setting that does not distinguish between infections occurring before or after *T* − *l* as (details in Supplementary Eqs. S13-S22)

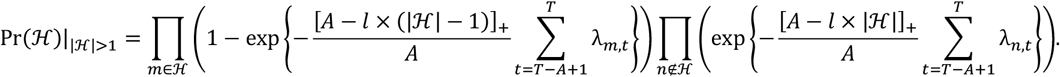

### General framework of titre-incorporated cross-reactive model

Let *Y*_*i*_ = {*Y*_*i*,D1_, *Y*_*i*,D2_, *Y*_*i*,D3_, *Y*_*i*,D4_} refer to the antibody titre level for individual *i*, where *Y*_*i*,*S*_ is the titre level against serotype *S*. For individuals with the number of historical infections |ℋ|, we define the probability density function (pdf) of *Y*_*i*,*S*_ as 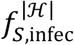 if *S* is one of the infecting serotypes, 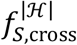 if individual *i* is unexposed to *S* and has positive cross-reactive antibodies against *S*, 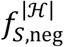 if individual *i* is unexposed to *S* and has negative antibodies against *S*. The likelihood of serostatus observation *T*_*i*_ = τ_*i*_ and titre levels *Y*_*i*_ = *y*_*i*_ is expanded based on Model (1) as below.

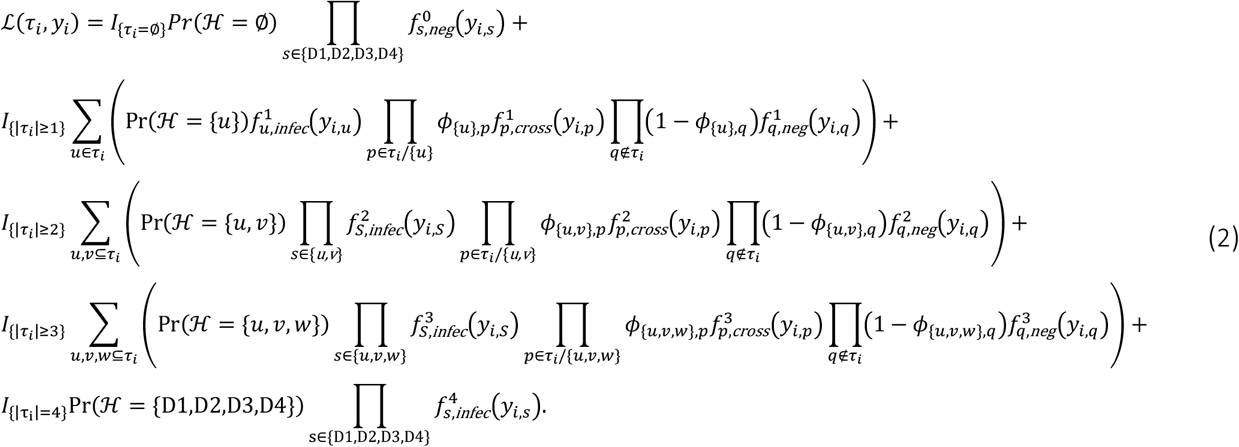

In this study, we specify Pr(ℋ) in the titre-incorporated model (Eq.(2)) within a unified framework as in the serostatus-based model (Eq.(1)). We transformed raw titre values as *Y*_*i*,*s*)_ = log 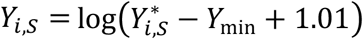where 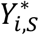 is the untransformed value, *Y*_min_ = 3.32 is the minimum titre value recorded, adding offset 1.01 is to constrain *Y*_*i*,*s*_ > 0. The transformed titre values *Y*_*i*,*s*_ range from 0.01 to 2.08. We use gamma distributions for *y*_*i*,*s*_ and 2.081 − *y*_5,)_ to capture titre values concentrated near the lower detection limit and assay-level maximum, respectively. Intermediate titres were modelled using normal distribution. Titre distributions were parameterized using serotype-specific baseline values for the fully-naïve state, additive infection-number effects, and response-specific boosts. Infection-number effects were modelled as separate non-negative increments for exposed and unexposed serotypes. Response-specific boosts were estimated separately for infecting and cross-reactive serotypes, with parameters varying by serotype. Titres to exposed serotypes were capped at the assay maximum after the third infection.

We also allow for two additional sources of titre heterogeneity. First, we include a proportion of individuals with elevated dengue antibody titres attributable to exposure to other pathogens. The component is assumed to be distinguishable in individuals naïve to 4 dengue serotypes or with a prior primary dengue infection. Second, after primary infections, we consider the possible scenario that individuals remain seronegative to heterotypic serotypes but carry low-level cross-reactive antibodies, with titre increases insufficient to exceed the seropositivity threshold. Detailed constructions of 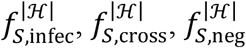 are available in Supplementary Eqs. S23-S24.

### Model fitting and posterior estimation

The models were implemented in Bayesian framework using *Rstan* package (*20*), running 8 Hamiltonian Monte Carlo (HMC) chains with 4,000 iterations each, including 500 warm-up iterations per chain. In serostatus-based model, the log likelihood was calculated as ∑ *N*_*τ*,*A*,*T*,*C*_ log ℒ (*τ*) (*N*_*τ*,*A*,*Y*,*C*_: number of samples with serostatus *τ* from individuals aged *A* during year *T*, and collected in year *T* at city *C* ∈ {HCMC, KH}), where ℒ(*τ*) was constructed under Model (1) framework. The log likelihood calculation adjusted for subset-tested data in the simulation study is as in Supplementary Eq. S25. In titre-incorporated model, the log-likelihood was calculated as ∑_*i*_ log ℒ (*τ*_*i*_, *y*_*i*_), where ℒ(τ_*i*_, *y*_*i*_) was constructed under Model (2) framework. Informative prior was given to cross-protection period (*l*) according to the literature, which was normal distribution with mean 1.8 years and standard deviation 0.15 (*21, 22*). Uninformative or weakly informative priors were given to other parameters (Supplementary Eqs. S26-S27). Model convergence was assessed by the minimum effective sample size, maximum R hat value, and HMC trace plots (Supplementary Figures. S23-S24). Model fit was evaluated by comparing the model estimates of seroprevalence and titre distributions with the corresponding empirical values calculated from the data (Supplementary Figures. S25-S27). The sensitivity of model estimates to the informative prior on the cross-protection period was evaluated. Based on the model results, we further derived posterior estimates of seroprevalence, population immunity and susceptibility profiles, and individual-level infection histories (Supplementary Eqs. S28-S29).

## Data Availability

All data and code used in this study are available at https://github.com/chenyn226/dengue-cross-reactivity.

## Acknowledgments

We would like to thank Dr. Ziyuan Niu from National University of Singapore for her help proofreading the manuscript prior to submission and for her valuable suggestions that improved the writing quality.

## Author contributions

Conceptualization: Y.C., H.C.; Methodology: Y.C., H.C.; Software: Y.C.; Validation: Y.C., A.T.H., H.C.; Formal analysis: Y.C.; Investigation: Y.C.; Resources: H.C.; Data Curation: Y.C.; Visualization: Y.C., A.T.H., H.C.; Supervision: H.C.; Project administration: H.C.; Funding acquisition: A.T.H., H.C.; Writing - Original Draft: Y.C.; Writing - Review & Editing: Y.C., A.T.H., H.C.

## Data and materials availability

The data and code used in this study are available at https://github.com/chenyn226/dengue-cross-reactivity.

## References

1. World Health Organization, Dengue - Global situation (2023). https://www.who.int/emergencies/disease-outbreak-news/item/2023-DON498.

2. A. Wilder-Smith, E.-E. Ooi, O. Horstick, B. Wills Dengue. The Lancet 393, 350–363 (2019).

3. W. Dejnirattisai, A. Jumnainsong, N. Onsirisakul, P. Fitton, S. Vasanawathana, W. Limpitikul, C. Puttikhunt, C. Edwards, T. Duangchinda, S. Supasa, K. Chawansuntati, P. Malasit, J. Mongkolsapaya, G. Screaton, Cross-Reacting Antibodies Enhance Dengue Virus Infection in Humans. Science 328, 745–748 (2010).

4. A. L. St. John, A. P. S. Rathore, Adaptive immune responses to primary and secondary dengue virus infections. Nat. Rev. Immunol. 19, 218–230 (2019).

5. E. Finch, C. Chang, A. Kucharski, S. Sim, L.-C. Ng, R. Lowe, Climate variation and serotype competition drive dengue outbreak dynamics in Singapore. Nat. Commun. 16, 11364 (2025).

6. S. Biswal, H. Reynales, X. Saez-Llorens, P. Lopez, C. Borja-Tabora, P. Kosalaraksa, C. Sirivichayakul, V. Watanaveeradej, L. Rivera, F. Espinoza, L. Fernando, R. Dietze, K. Luz, R. Venâncio Da Cunha, J. Jimeno, E. López-Medina, A. Borkowski, M. Brose, M. Rauscher, I. LeFevre, S. Bizjajeva, L. Bravo, D. Wallace, Efficacy of a Tetravalent Dengue Vaccine in Healthy Children and Adolescents. N. Engl. J. Med. 381, 2009–2019 (2019).

7. H. E. Clapham, I. Rodriguez-Barraquer, A. S. Azman, B. M. Althouse, H. Salje, R. V. Gibbons, A. L. Rothman, R. G. Jarman, A. Nisalak, B. Thaisomboonsuk, S. Kalayanarooj, S. Nimmannitya, D. W. Vaughn, S. Green, I.-K. Yoon, D. A. T. Cummings, Dengue Virus (DENV) Neutralizing Antibody Kinetics in Children After Symptomatic Primary and Postprimary DENV Infection. J. Infect. Dis. 213, 1428–1435 (2016).

8. T. T. N. Thao, E. De Bruin, H. T. Phuong, N. H. Thao Vy, H.-J. Van Den Ham, B. A. Wills, N. T. H. Tien, H. T. Le Duyen, D. T. Trung, S. S. Whitehead, M. F. Boni, M. Koopmans, H. E. Clapham, Using NS1 Flavivirus Protein Microarray to Infer Past Infecting Dengue Virus Serotype and Number of Past Dengue Virus Infections in Vietnamese Individuals. J. Infect. Dis. 223, 2053–2061 (2021).

9. M. O’Driscoll, N. Hozé, N. Lefrancq, G. Ribeiro Dos Santos, D. Hoinard, M. Z. Rahman, K. K. Paul, A. M. Naser Titu, M. S. Alam, M. E. Hossain, J. Vanhomwegen, S. Cauchemez, E. S. Gurley, H. Salje, Epidemiological and antigenic inferences from serological cross-reactivity among arboviruses. Sci. Transl. Med. 17, eads8680 (2025).

10. H. T. Phuong, N. H. T. Vy, N. T. L. Thanh, M. Tan, E. De Bruin, M. Koopmans, M. F. Boni, H. E. Clapham, Estimating the force of infection of four dengue serotypes from serological studies in two regions of Vietnam. PLoS Negl. Trop. Dis. 18, e0012568 (2024).

11. United Nations Statistics Division, Population by age, sex and urban/rural residence (2022).

12. D.-T. Chu, V. T. N. Ngoc, Y. Tao, Zika virus infection in Vietnam: current epidemic, strain origin, spreading risk, and perspective. Eur. J. Clin. Microbiol. Infect. Dis. 36, 2041–2042 (2017).

13. S.-L. Low, S. Lam, W.-Y. Wong, D. Teo, L.-C. Ng, L.-K. Tan, Dengue Seroprevalence of Healthy Adults in Singapore: Serosurvey Among Blood Donors, 2009. Am. Soc. Trop. Med. Hyg. 93, 40–45 (2015).

14. L. K. Tan, S. L. Low, H. Sun, Y. Shi, L. Liu, S. Lam, H. H. Tan, L. W. Ang, W. Y. Wong, R. Chua, D. Teo, L. C. Ng, A. R. Cook, Force of Infection and True Infection Rate of Dengue in Singapore: Implications for Dengue Control and Management. Am. J. Epidemiol. 188, 1529–1538 (2019).

15. A.-F. Taurel, C. Q. Luong, T. T. T. Nguyen, K. Q. Do, T. H. Diep, T. V. Nguyen, M. T. Cao, T. N. D. Hoang, P. T. Huynh, T. K. L. Huynh, M. H. Le, J. Nealon, A. Moureau, Age distribution of dengue cases in southern Vietnam from 2000 to 2015. PLoS Negl. Trop. Dis. 17, e0011137 (2023).

16. B. Labrosse, M. Tourdjman, R. Porcher, J. LeGoff, X. De Lamballerie, F. Simon, J.-M. Molina, F. Clavel, Detection of Extensive Cross-Neutralization between Pandemic and Seasonal A/H1N1 Influenza Viruses Using a Pseudotype Neutralization Assay. PLoS ONE 5, e11036 (2010).

17. L. C. Katzelnick, J. M. Fonville, G. D. Gromowski, J. B. Arriaga, A. Green, S. L. James, L. Lau, M. Montoya, C. Wang, L. A. VanBlargan, Dengue viruses cluster antigenically but not as discrete serotypes. Science 349, 1338–1343 (2015).

18. L. C. Katzelnick, A. Coello Escoto, A. T. Huang, B. Garcia-Carreras, N. Chowdhury, I. Maljkovic Berry, C. Chavez, P. Buchy, V. Duong, P. Dussart, G. Gromowski, L. Macareo, B. Thaisomboonsuk, S. Fernandez, D. J. Smith, R. Jarman, S. S. Whitehead, H. Salje, D. A. T. Cummings, Antigenic evolution of dengue viruses over 20 years. Science 374, 999–1004 (2021).

19. K. T. D. Thai, T. Q. Binh, P. T. Giao, H. L. Phuong, L. Q. Hung, N. V. Nam, T. T. Nga, J. Groen, N. Nagelkerke, P. J. De Vries, Seroprevalence of dengue antibodies, annual incidence and risk factors among children in southern Vietnam. Trop. Med. Int. Health 10, 379–386 (2005).

20. B. Carpenter, A. Gelman, M. D. Hoffman, D. Lee, B. Goodrich, M. Betancourt, M. A. Brubaker, J. Guo, P. Li, A. Riddell, Stan: A probabilistic programming language. J. Stat. Softw. 76 (2017).

21. N. G. Reich, S. Shrestha, A. A. King, P. Rohani, J. Lessler, S. Kalayanarooj, I.-K. Yoon, R. V. Gibbons, D. S. Burke, D. A. T. Cummings, Interactions between serotypes of dengue highlight epidemiological impact of cross-immunity. J. R. Soc. Interface 10, 20130414 (2013).

22. M. Montoya, L. Gresh, J. C. Mercado, K. L. Williams, M. J. Vargas, G. Gutierrez, G. Kuan Gordon, A. Balmaseda, E. Harris, Symptomatic Versus Inapparent Outcome in Repeat Dengue Virus Infections Is Influenced by the Time Interval between Infections and Study Year. PLoS Negl. Trop. Dis. 7, e2357 (2013).

